# Toward point-of-care diagnostics of *Candida auris*

**DOI:** 10.1101/2020.02.10.20021683

**Authors:** Geoffrey Mulberry, Sudha Chaturvedi, Vishnu Chaturvedi, Brian N. Kim

## Abstract

*Candida auris* is a multidrug-resistant yeast that presents global health threat for the hospitalized patients. Early diagnostic of *C. auris* is crucial in control, prevention, and treatment. *Candida auris* is difficult to identify with standard laboratory methods and often can be misidentified leading to inappropriate management. A newly-devised real-time PCR assay played an important role in the ongoing investigation of the *C. auris* outbreak in New York metropolitan area. The assay can rapidly detect *C. auris* DNA in surveillance and clinical samples with high sensitivity and specificity, and also useful for confirmation of *C. auris* cultures. Despite its positive impact, the real-time PCR assay is difficult to deploy at frontline laboratories due to high-complexity set-up and operation. Using a low-cost handheld real-time PCR device, we show that the *C. auris* can potentially be identified in a low-complexity assay without the need for high-cost equipment. An implementation of low-cost real-time PCR device in hospitals and healthcare facilities is likely to accelerate the diagnosis of *C. auris* and for control of the global epidemic.

## I. Introduction

*Candida auris* is an emerging multidrug-resistant yeast, which presents a serious global health threat. The largest, localized, uninterrupted *C. auris* outbreak in the US healthcare facilities continues to afflict the New York metro areas (2013-2019) [1], [2]. The New York State Department of Health (NYSDOH) Mycology Laboratory has been serving a crucial role in developing rapid, and high-throughput assays that are sensitive and specific to *C. auris*, and in processing over 20,000 clinical samples in the state for diagnosis and surveillance purposes [3], [4]. Diagnosing *C. auris* infections in the early stage is crucial in the administration of empirical antifungal therapy and the implementation of infection control measures. Many early-stage diagnostic devices have been developed in recent years to detect various pathogens including *C. auris* with improved sensitivity and cheaper devices [5]–[10], but they fail to improve the medical logistical system for early diagnostics, due to the labor-intensive sample preparation process [8], [11]– [16].

Wadsworth Center NYSDOH’s *Candida auris* LDT, standard operating procedures, and validation results were shared extensively with NY clinical laboratories, Centers for Disease Control, and other public health laboratories. Several publications have described other approaches for real-time PCR confirmation of *C. auris* [17]–[19]. However, the adoption of LDT technologies is progressing slowly while the affected facilities and sample numbers continue to grow. In response, the US Food and Drug Administration (FDA) permitted marketing for a new use of the BRUKER MALDI Biotyper CA system for *C. auris* isolates [20]. However, isolates are not available early in the clinical and epidemiological investigations. The scope and complexity of this outbreak requires most hospital laboratories to perform a *C. auris* PCR screening test for faster implementation of the infection prevention measures, while more complex testing for fungal culture, antifungal testing, and genotyping continues at the reference laboratories.

The Centers for Disease Control and Prevention (CDC) recommends using real-time PCR for the identification of *C. auris* to obtain a diagnosis and to determine the proper treatment [21]. Real-time PCR diagnostic tests are especially attractive as they reveal both the presence and quantity of the pathogen with high sensitivity and specificity with rapid turnaround. However, the implementation of real-time PCR is challenging for many frontline laboratories due to the process’ complexities and high cost (>$30,000), and thus is only located in a centralized laboratory. These barriers are especially prohibitive for the point-of-care diagnostics of infectious diseases. Therefore, the integration of efficiency, portability, and effectiveness has been identified as a key priority for the development of novel, point-of-care devices. Alternative techniques with a more accessible nature, such as rapid diagnostic tests (RDT), are often used to make a diagnosis at the cost of inaccuracy. The real-time PCR method is a robust and reliable technique that amplifies the target DNA sequence while monitoring a signal originating from intercalating probes to quantify the amount of DNA being amplified. Real-time PCR can be performed with custom-designed primers to provide a highly specific diagnosis by amplifying a specific sequence of DNA. This enables multiplexed detection with the capability of differentiating various types of *Candida* to coordinate proper treatment.

## II. Point-of-care real-time PCR Device

The high cost of real-time PCR machines has limited their accessibility to a centralized laboratory resulting in a long turnaround for diagnostics. We recently developed a set of 3D manufacturing methods for fabrication of a portable real-time PCR device [22] (Fig. 1a-c). The key advantage of this approach is the ability to upload the digital format of the design files to the internet for wide distribution, which people at any location can simply download and feed into their 3D printers for rapid manufacturing. The portable real-time PCR device designed using these methods is battery-operated, with a size of 12 × 7 × 6 cm^3^ and weight of only 214 g. The production cost of the real-time PCR device was ∼300 USD. The entire process of thermal cycling and time-coordinated fluorescence reading is automated by closed-loop feedback. The 3D-manufactured real-time PCR device was validated by detection of lentivirus, as a proxy for medically important retroviruses such as HIV. The C^t^ was measured from fluorescence recordings (Fig. 1d). The device can operate for hours without any external power and demonstrated robust detection and quantitation of viral particles across a range of clinically relevant concentrations [22] (Fig. 1d). Adaptation and refinement of this device for real-time PCR detection of *C. auris* may result in an innovative point-of-care system for infectious disease diagnosis.

**Figure 1.**
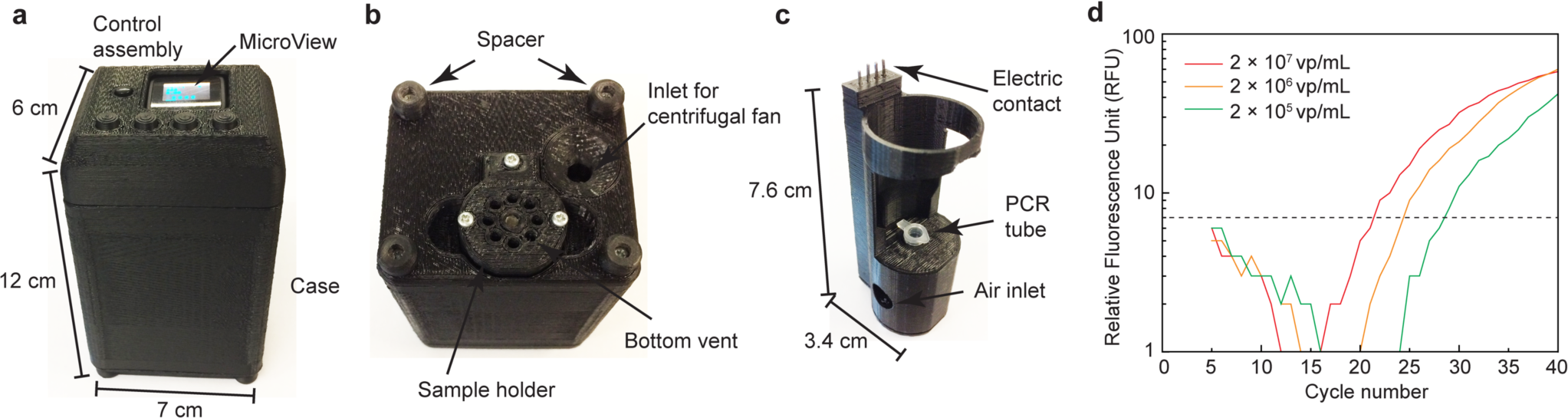
3D manufactured real-time PCR device for rRT-PCR-based virus detection. (a) The amplification and fluorescence reading status is displayed in real time. (b) The bottom view showing an air inlet for the centrifugal fan. (c) The sample holder holds the PCR tube. (d) rRT-PCR Fluorescence readings show the shift in the intensity measurements corresponding to the differing concentrations of virus. Reprint from [22].

## III. Detecting *C. auris* using the Portable Device

We used the purified *C. auris* DNA to test the portable device’s ability to detect the presence of *C. auris*. This is a proof-of-principle experiment to show that *C. auris* can potentially be detected with a low-cost device at a primary care. The TaqMan-based assay reagents and probes, developed at the Wadsworth Center NYSDOH that is sensitive and specific to *C. auris*, is used in the low-cost real-time PCR device (Fig. 2). The user interface allows the user to change variables associated with real-time PCR cycles, such as thermocycling temperatures, durations, and fluorescence reading (Fig. 2c). In the control assembly, an LCD screen will display the user’s selections that can be changed by the four navigation buttons. During PCR, the LCD displays the temperature to indicate the progress of the assay and show fluorescence readings. The resulting flouresence data revealed that this device is indeed capable of detecting the presence of *C. auris* gene in the sample in less than 2 hours (Fig. 2b).

**Figure 2.**
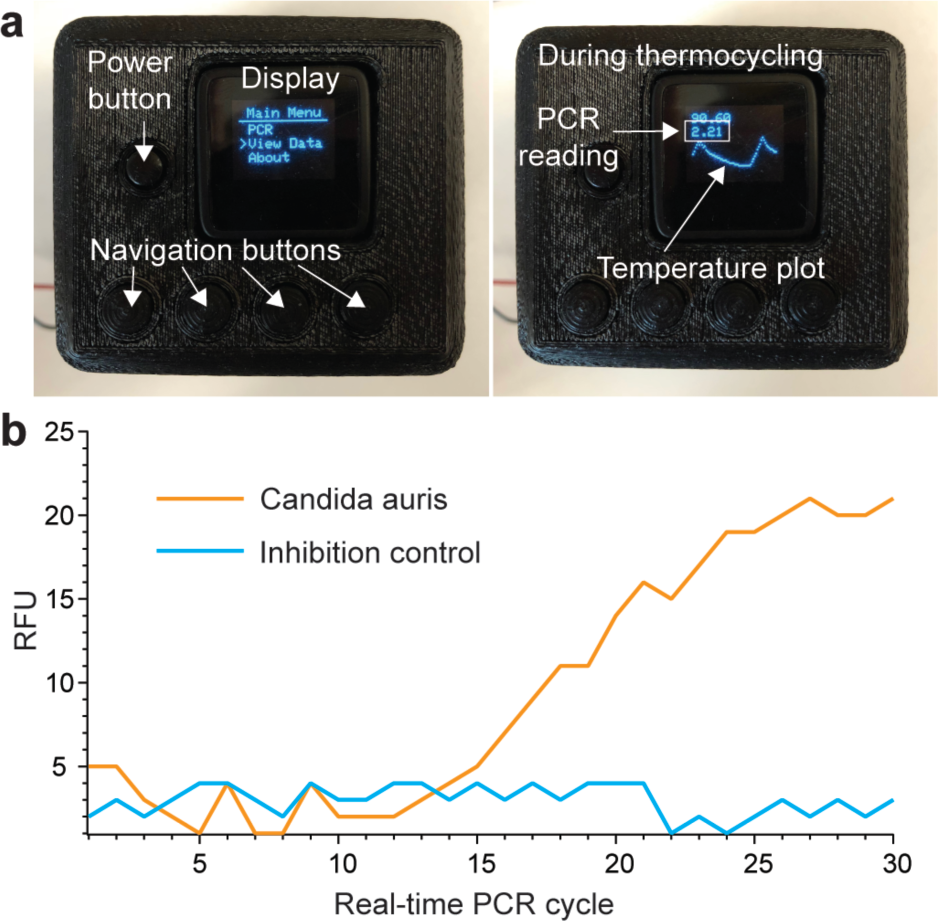
Adaption of 3D manufactured real-time PCR device for *C. auris* detection. (a) 3D assembly of the two-tube real-time PCR system. Fluorescence measurements of the DNA amplification are enable using integrated filter sets, LEDs, and photodiodes. Control assembly contains the user interface, including navigation buttons, an LCD display, and a power button. (d) Purified C. auris DNA detection using the handheld real-time PCR device.

## IV. Conclusion

In this work, we discuss the need of rapid point-of-care diagnostics for the detection *C. auris* to prevent further spread, and reduce the mortality by early detection. Using the low-cost, handheld real-time PCR device, we are able to reveal the presence of *C. auris* DNA in the sample. Potentially, these type of low-cost nucleic-acid testing devices can facilitate the wide use of real-time PCR for the early detection of *C. auris*. It is important to note that the major challenges facing point-of-care diagnostic also includes the sample preparation which this work does not address. The sample collected from the patient requires a sample preparation step where the DNA is purified and extracted from the sample before they can be used for real-time PCR detection. This preparation step is labor intensive and requires specialized skill, therefore imposes a challenge in wide use of real-time PCR assay in frontline laboratories and clinics. We are working toward simplifying and automating this step to enable the application of low-cost real-time PCR devices in frontline laboratories and clinics [23].

## Data Availability

The raw data will be available upon request.

## References

[1] E. Adams, M. Quinn, S. Tsay, E. Poirot, S. Chaturvedi, K. Southwick, J. Greenko, R. Fernandez, A. Kallen, S. Vallabhaneni, Haley, B. Hutton, D. Blog, E. Lutterloh, H. Zucker, C. Bucher, R. L. Erazo, R. Giardina, J. Glowicz et al., “Candida auris in healthcare facilities, New York, USA, 2013–2017,” Emerg. Infect. Dis., vol. 24, no. 10, pp. 1816–1824, Oct. 2018.

[2] Y. Zhu, B. O’Brien, L. Leach, A. Clark, M. Bates, E. Adams, B. Ostrowsky, M. Quinn, E. Dufort, K. Southwick, R. Erazo, V. B. Haley, C. Bucher, V. Chaturvedi, R. J. Limberger, D. Blog, E. Lutterloh and S. Chaturvedi, “Laboratory Analysis of an Outbreak of Candida auris in New York from 2016 to 2018-Impact and Lessons Learned,” J. Clin. Microbiol., Dec. 2019.

[3] L. Leach, Y. Zhu and S. Chaturvedi, “Development and Validation of a Real-Time PCR Assay for Rapid Detection of Candida auris from Surveillance Samples,” J. Clin. Microbiol., vol. 56, no. 2, Feb. 2018.

[4] L. Leach, A. Russell, Y. Zhu, S. Chaturvedi and V. Chaturvedia, “A rapid and automated sample-to-result Candida auris real-time pcr assay for high-throughput testing of surveillance samples with the bd max open system,” J. Clin. Microbiol., vol. 57, no. 10, Oct. 2019.

[5] A. Moody, “Rapid diagnostic tests for malaria parasites.,” Clin. Microbiol. Rev., vol. 15, no. 1, pp. 66–78, Jan. 2002.

[6] N. E. Rosenberg, G. Kamanga, S. Phiri, D. Nsona, A. Pettifor, S. E. Rutstein, D. Kamwendo, I. F. Hoffman, M. Keating, L. B. Brown, B. Ndalama, S. A. Fiscus, S. Congdon, M. S. Cohen and W. C. Miller, “Detection of acute HIV infection: a field evaluation of the determine® HIV-1/2 Ag/Ab combo test.,” J. Infect. Dis., vol. 205, no. 4, pp. 528–34, Feb. 2012.

[7] T. Hänscheid, M. Rebelo and M. P. Grobusch, “Point-of-care tests: where is the point?,” Lancet. Infect. Dis., vol. 14, no. 10, p. 922, Oct. 2014.

[8] A. St John and C. P. Price, “Existing and Emerging Technologies for Point-of-Care Testing.,” Clin. Biochem. Rev., vol. 35, no. 3, pp. 155–67, Aug. 2014.

[9] M. Sher, R. Zhuang, U. Demirci and W. Asghar, “Paper-based analytical devices for clinical diagnosis: recent advances in the fabrication techniques and sensing mechanisms.,” Expert Rev. Mol. Diagn., vol. 17, no. 4, pp. 351–366, 2017.

[10] C. Wongsrichanalai, M. J. Barcus, S. Muth, A. Sutamihardja and W. H. Wernsdorfer, “A review of malaria diagnostic tools: microscopy and rapid diagnostic test (RDT).,” Am. J. Trop. Med. Hyg., vol. 77, no. 6 Suppl, pp. 119–27, Dec. 2007.

[11] N. T. Ho, A. Fan, C. M. Klapperich and M. Cabodi, “Sample concentration and purification for point-of-care diagnostics.,” Conf. Proc Annu. Int. Conf. IEEE Eng. Med. Biol. Soc. IEEE Eng. Med. Biol. Soc. Annu. Conf., vol. 2012, pp. 2396–9, 2012.

[12] J. Pang, P. Y. Chia, D. C. Lye and Y. S. Leo, “Progress and Challenges towards Point-of-Care Diagnostic Development for Dengue.,” J. Clin. Microbiol., vol. 55, no. 12, pp. 3339–3349, Dec. 2017.

[13] T. R. Kozel and A. R. Burnham-Marusich, “Point-of-Care Testing for Infectious Diseases: Past, Present, and Future.,” J. Clin. Microbiol., vol. 55, no. 8, pp. 2313–2320, Aug. 2017.

[14] N. Ali, R. de C. P. Rampazzo, A. D. T. Costa and M. A. Krieger, “Current Nucleic Acid Extraction Methods and Their Implications to Point-of-Care Diagnostics,” Biomed Res. Int., vol. 2017, pp. 1–13, Jul. 2017.

[15] C. D. Chin, S. Y. Chin, T. Laksanasopin and S. K. Sia, “Low-Cost Microdevices for Point-of-Care Testing,” Springer, Berlin, Heidelberg, 2013, pp. 3–21.

[16] D. Mabey, R. W. Peeling, A. Ustianowski and M. D. Perkins, “Diagnostics for the developing world,” Nat. Rev. Microbiol., vol. 2, no. 3, pp. 231–240, Mar. 2004.

[17] M. Kordalewska, Y. Zhao, S. R. Lockhart, A. Chowdhary, I. Berrio and D. S. Perlin, “Rapid and accurate molecular identification of the emerging multidrug-resistant pathogen Candida auris,” J. Clin. Microbiol., vol. 55, no. 8, pp. 2445–2452, Aug. 2017.

[18] A. Ahmad, J. E. Spencer, S. R. Lockhart, S. Singleton, D. J. Petway, D. A. Bagarozzi and O. T. Herzegh, “A high-throughput and rapid method for accurate identification of emerging multidrug-resistant Candida auris,” Mycoses, vol. 62, no. 6, pp. 513–518, Jun. 2019.

[19] A. Arastehfar, W. Fang, F. Daneshnia, A. M. S. Al-Hatmi, W. Liao, W. Pan, Z. Khan, S. Ahmad, K. Rosam, M. Lackner, C. Lass-Flörl, F. Hagen and T. Boekhout, “Novel multiplex real-time quantitative PCR detecting system approach for direct detection of Candida auris and its relatives in spiked serum samples,” Future Microbiol., vol. 14, no. 1, pp. 33–45, Jan. 2019.

[20] J. R. Bao, R. N. Master, K. N. Azad, D. A. Schwab, R. B. Clark, R. S. Jones, E. C. Moore and K. L. Shier, “Rapid, accurate identification of candida auris by using a novel matrix-assisted laser desorption ionization–time of flight mass spectrometry (MALDI-TOF MS) database (library),” Journal of Clinical Microbiology, vol. 56, no. 4. American Society for Microbiology, 01-Apr-2018.

[21] C. for Disease Control, “REAL-TIME PCR BASED IDENTIFICATION OF CANDIDA AURIS USING APPLIED BIOSYSTEMS 7500 FAST REAL-TIME PCR PLATFORM.”

[22] G. Mulberry, K. A. White, M. Vaidya, K. Sugaya and B. N. Kim, “3D printing and milling a real-time PCR device for infectious disease diagnostics,” PLoS One, vol. 12, no. 6, pp. 1–18, 2017.

[23] G. Mulberry, A. Vuillier, M. Vaidya, K. Sugaya and B. N. Kim, “Handheld battery-operated sample preparation device for qPCR nucleic acid detections using simple contactless pouring,” Anal. Methods, vol. 10, no. 38, pp. 4671–4679, Oct. 2018.

